# AFFORDABILITY OF INTOXICATION FROM CHEAP ETHANOL: EVIDENCE FROM RETAIL ALCOHOL MARKETS IN UGANDA

**DOI:** 10.64898/2026.06.18.26356010

**Authors:** Muhamudu Tumwine, Kennedy Niwagaba, Simon Peter Rwakahangi, Betty Kwagala, Ted R Miller, Cosmas Zyambo, Angela Rizzo, Tom Achoki

## Abstract

**Background:** Alcohol affordability is a determinant of consumption and alcohol-related harm. In many low- and middle-income countries (LMICs), informal production, variable alcohol strength, and non-standard packaging complicate conventional affordability measures, limiting evidence on the economic accessibility of alcohol and the cost of intoxication.

**Objective:** To assess the affordability of intoxication in Uganda by estimating the cost of obtaining ethanol to reach intoxication across alcohol products, packaging types, and retail contexts.

**Methods:** Data were collected on 824 alcoholic beverages from urban, rural, and urban-slum retail markets. Ethanol-standardized pricing (price per gram of alcohol) was calculated, and the cost of consuming 60 g of ethanol was estimated. Multivariate regression identified determinants of ethanol affordability.

**Results:** Affordability varied by product type and packaging. Opaque beers and illicit spirits provided the cheapest pathways to intoxication, with median costs of UGX 1,200–1,500 per 60 g of ethanol. Plastic packaging was associated with lower ethanol costs than glass packaging. Ethanol prices differed across formal and informal markets (p < 0.01), while rural areas and urban informal settlements had 20–25% lower costs than urban areas. Regulatory status alone did not predict affordability.

**Conclusions:** In Uganda’s diverse alcohol market, affordability is driven by access to ethanol rather than beverage price alone. Low-cost, high-strength alcohol sold through informal channels enables intoxication at minimal expense, among disadvantaged populations.

**Implications:** Alcohol policies should target ethanol content through minimum unit pricing, alcohol-content-based taxation, and regulation of informal markets and packaging practices to reduce harmful consumption and inequities.

## Introduction

Public health responses to alcohol-related harm have increasingly emphasized the role of pricing policies in shaping consumption patterns (1). A substantial body of evidence demonstrates that alcohol affordability, defined as the relationship between price and purchasing power (2) is a key determinant of population-level drinking and associated harms (1). Policies such as excise taxation and minimum unit pricing are therefore designed to increase the economic cost of alcohol, particularly for high-strength products, thereby reducing harmful consumption (3–5). Systematic reviews further confirm that increases in alcohol prices are consistently associated with reductions in consumption and alcohol-attributable harm(6–8). These effects operate through affordability, which has become a central construct in alcohol policy research (9–11).

However, conventional approaches to measuring affordability may be conceptually limited. Much of the existing evidence is derived from high-income settings characterized by formalized markets, standardized products, and regulated pricing structures. In these contexts, alcohol is typically sold in uniform volumes with clearly labelled alcohol content, enabling consistent measurement of price and affordability (2,12,13). These assumptions do not hold in many low- and middle-income countries (LMICs), where alcohol markets are fragmented and heterogeneous. Beverages are often sold in non-standard quantities, alcohol content varies widely within and across product categories, and pricing is frequently determined at the point of sale (14). Under such conditions, conventional affordability measures such as price per beverage or expenditure relative to income may provide an incomplete representation of actual alcohol exposure (15,16).

A growing body of research further shows that alcohol markets in LMICs are shaped by the coexistence of formal and informal systems, alongside the expansion of commercial actors into less regulated environments (17,18). In East Africa, these dynamics are layered onto long-standing traditions of local alcohol production and distribution, which continue to influence contemporary consumption patterns and market structures (19). The result is a diverse alcohol environment in which price signals alone may not accurately reflect access to ethanol.

From a public health perspective, it is the quantity of ethanol consumed rather than the volume of beverage that determines intoxication and risk of harm (20). This has led to increasing emphasis on ethanol-standardized measures, such as price per gram of alcohol, which allow for meaningful comparisons across beverages with differing strengths (21,22). Closely related is the concept of the cost of intoxication, defined as the minimum financial outlay required to consume a given quantity of ethanol sufficient to induce intoxication (23). This metric shifts the analytical focus from beverage consumption to economic accessibility of intoxication an underexplored dimension of alcohol availability. Yet empirical applications of these approaches remain limited in LMIC settings, particularly within informal retail markets where both alcohol content and pricing are highly variable.

Uganda presents a particularly relevant context for examining these dynamics. According to World Health Organisation (WHO) and Uganda Policy Alliance (UAPA), Uganda ranks first in Africa, exceeding the continent’s average of 4.5-6 liters of pure alcohol per person per year and the global average of ∼6.18 liters per person yearly (24,25). The country bears a substantial burden of alcohol-related morbidity and mortality (26–28), and alcohol is widely available across formal outlets, informal vendors, and small-scale producers. Products vary considerably in alcohol concentration, packaging, and pricing structure, with high-strength spirits often sold in small, low-cost units that reduce the immediate financial barrier to consumption. These characteristics suggest that the effective price of ethanol and, by extension, the cost of intoxication may differ substantially across products and market segments.

While previous studies in Uganda have examined alcohol consumption patterns, health risks, and characteristics of informal alcohol markets (29–31), research is scarce on the economic structuring of ethanol access. In particular, limited evidence exists on how variations in alcohol strength and retail pricing translate into the affordability of intoxication. Moreover, the extent to which product characteristics, alcohol concentration, and retail context independently shape ethanol pricing has not been systematically examined using multivariate approaches.

This study addresses these gaps by examining the affordability of intoxication within retail alcohol markets in Uganda. Using detailed data on product characteristics, ethanol concentration, and retail prices, the study derives standardized measures of price per gram of ethanol and estimates the cost of intoxication across beverage types. It further compares formal and informal market segments and applies multivariate regression to identify the structural determinants of ethanol affordability. Focusing on the economic pathways through which intoxication is achieved, the study provides evidence to inform alcohol pricing policies and regulatory interventions in LMIC settings.

## Methods

### Ethics statement

Ethical approval for the study was granted by the Uganda National Council for Science and Technology (UNCST) and the Makerere University School of Social Sciences Research Ethics Committee (MAKSSREC) (Ref. No. MAKSSREC2024-782). Additional administrative clearance was obtained from the Kampala Capital City Authority (KCCA) (Ref. No. DPHE/KCCA/130/01).

### Study Design

This study employed a cross-sectional analytical design to examine the affordability of ethanol and the cost of intoxication in Uganda’s retail alcohol markets. The design was informed by public health and alcohol policy frameworks, emphasizing alcohol availability, price, and strength as key determinants of consumption and harm (4,20,28). Rather than focusing solely on beverage price, the study applied an ethanol-standardized approach, comparing alcoholic drinks based on the cost of pure alcohol and the financial outlay required to achieve an intoxicating dose (8,23).

### Study Setting and Population

Data was collected across diverse retail alcohol markets, including urban, urban slum, and rural settings across the western, eastern, northern, and central regions of Uganda. The unit of observation was the retail alcohol product available for sale, with each unique combination of brand, declared ABV, packaging size, and container type treated as a distinct observation. The study included 824 beverages: 104 clear beers, 31 opaque beers, 478 spirits, and 211 wines, consistent with WHO classifications(32,33). Retail outlets such as bars, shops, and kiosks served as the site of observation, while wholesalers and distributors were excluded to avoid bulk price distortions. Alcohol drinks without declared ABV% were not included as part of this study.

### Sampling and Data Collection

Retail outlets were sampled using a systematic walk-through approach along predefined commercial routes. A purposive strategy ensured representation across urban (n = 399), urban informal (n = 185), and rural (n = 240) areas. Both licensed and unlicensed outlets were included to capture formal and informal market structures. Within each outlet, all beverages were recorded, with product entries distinguished by differences in ABV, packaging, or container type (18,34).

Primary data collected included retail price (UGX), beverage volume (ml), ABV (%), packaging characteristics, and regulatory indicators (e.g., certification, tax stamps). Secondary data from the Uganda Revenue Authority, Uganda National Bureau of Standards, and the Ministry of Health contextualized taxation and regulatory frameworks.

### Validity and Reliability

Enumerators received standardized training, and data collection procedures were pilot-tested. Inter-rater reliability was assessed via duplicate pilot-test recordings, yielding Cronbach’s α = 0.96 and Pearson r = 0.92, indicating high agreement.

### Variable and Measurements

#### Ethanol Content

The ethanol content of each beverage was calculated using standard conversion methods:

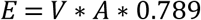

where E = grams of ethanol, V = volume (ml), A = alcohol by volume (proportion), and 0.789 = ethanol density (g/ml)

#### Primary Outcome: Cost of Ethanol

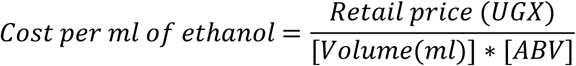

This measure standardized alcohol prices across beverages with different strengths and volumes, allowing direct comparison of ethanol affordability.

#### Derived Outcome: Cost of Intoxication

Affordability of intoxication was operationalized as the cost of obtaining 60 grams of ethanol, consistent with(35) definitions of heavy episodic drinking.

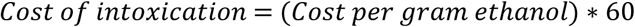

Where:

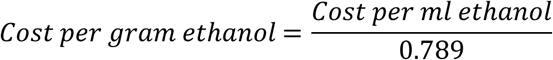

#### Independent Variables

Variables were grouped into:

- **Product:** Alcohol type (spirits, clear beer, opaque beer, wine), Country of origin (imported vs domestic), Packaging size (≤200 ml, 201–375 ml, 750 ml, >750 ml), Packaging material (glass, plastic, cans)
- **Market:** Retail outlet type (bar, shop, kiosk), Location (urban, urban informal, rural), Availability (daily, weekly, monthly)
- **Regulatory:** A product was determined as illicit if it scored more than half from the following indicators: absence of UNBS seal (Q Mark), untaxed, plastic packaging, packaged in small volumes of 200ml or below, and sold below Ugx 2000. From this, a composite variable - Illicitness was measured as a binary indicator where 1 was non-compliant, and 0 was compliant.

### Data Analysis

Data were cleaned, coded, and analysed using Stata version 18. Descriptive statistics summarized product characteristics, ethanol concentration, and retail prices. Cost per ml of ethanol and cost of intoxication were computed for all beverages and compared across formal and informal market segments.

Given right-skewed distributions of prices, unit cost variables were log-transformed. Non-parametric bootstrapping with 1,000 resamples estimated robust standard errors and confidence intervals.

A multiple linear regression model examined determinants of ethanol price, with log-transformed cost per ml ethanol as the dependent variable and independent variables including product type, packaging, outlet type, location, regulatory status, and market factors. Coefficients with p < 0.05 were considered statistically significant.

## Results

### Product Characteristics of Alcoholic Beverages in Retail Markets

This section describes the distribution of alcoholic beverages across product, packaging, and retail characteristics to contextualize market structure.

The results from the composite score show that of the different alcohol products commercially sold in the Ugandan retail market with declared ABV%, illicit alcohol products account for 27.6% of the total sample size. Notably, opaque beers account for the highest illicit products at 96.8% followed by spirits at 40.6%.

The final dataset comprised 824 alcoholic beverage observations collected from retail markets across Uganda (Table 1). The sample showed near uniform distribution between imported (48.3%) and domestically produced beverages (51.7%).

**Table 1:** Results from the Composite Score of Alcoholic Beverages.

| Alcohol Type | Illicit | Licit | Total |
| --- | --- | --- | --- |
| Clear Beer | 0 | 104(100%) | 104(100%) |
| Opaque Beer | 30(96.8%) | 1(3.2%) | 31(100%) |
| Spirits | 194(40.6%) | 284(59.4%) | 478(100%) |
| Wines | 1(0.5%) | 210(99.5%) | 211(100%) |
| Total | 227(27.6%) | 597(72.4%) | 824(100%) |

Geographically, nearly half of the products were obtained from urban areas (48.4%), with the remainder distributed across rural (29.1%) and urban informal settlements (22.5%). Retail distribution was concentrated in bars (59.7%), followed by shops (33.6%) and kiosks (6.7%).

In terms of product composition, spirits constituted the majority of the sample (58.0%), combining licit (34.5%) and illicit (23.5%) categories. Wines accounted for 25.6%, while clear beer (12.6%) and opaque beer (3.8%) represented smaller shares.

Packaging was predominantly in standard 750 ml bottles (50.0%), followed by smaller 200 ml units (29.4%). Glass bottles were the dominant packaging material (71.0%), with plastic containers (26.2%) and cans (2.8%) comprising smaller proportions.

In total, 72.5% of beverages were classified as licit, while 27.5% were illicit, indicating a substantial presence of informal alcohol within the retail market.

### Ethanol Concentration across Alcohol Products

Variation in alcohol strength (ABV) across beverage types determines the quantity of ethanol available per unit volume and is central to standardizing affordability comparisons.

Ethanol concentration varied across beverage categories (Table 2). Illicit spirits recorded the highest mean alcohol by volume (ABV) at 39.9%, followed by licit spirits at 37.6%. Wines (12.1%), opaque beers (10.1%), and clear beers (5.3%) exhibited lower alcohol content.

**Table 2:** Product Characteristics of Alcoholic Beverages.

| Variable | Freq. | Percent (%) |
| --- | --- | --- |
| **Country of Origin** |  |  |
| Imported | 398 | 48.3 |
| Uganda | 426 | 51.7 |
| **Location of Sale** |  |  |
| Urban | 399 | 48.4 |
| Urban Slum | 185 | 22.5 |
| Rural | 240 | 29.1 |
| **Alcohol Type** |  |  |
| Clear Beer | 104 | 12.6 |
| Opaque Beer | 31 | 3.8 |
| Licit Spirits | 284 | 34.5 |
| Wines | 211 | 25.6 |
| Illicit Spirits | 194 | 23.5 |
| **Retail Outlet Type** |  |  |
| Kiosk | 55 | 6.67 |
| Shop | 277 | 33.6 |
| Bar | 492 | 59.7 |
| **Packaging Size** |  |  |
| Half Pint (200ml) | 242 | 29.4 |
| Half Bottle (375ml) | 92 | 11.2 |
| Standard Bottle (750ml) | 410 | 50.0 |
| Litres | 80 | 9.7 |
| **Packaging Type** |  |  |
| Can | 23 | 2.8 |
| Plastic | 216 | 26.2 |
| Glass | 585 | 71 |
| **Illicitness** |  |  |
| Illicit | 227 | 27.6 |
| Licit | 597 | 72.5 |
| **Total** | **824** | **100** |

Variation within categories was also evident. Licit spirits showed the widest range (15.0%–47.4%), while illicit spirits ranged from 22.0% to 43.0%. Beer categories displayed narrower ranges but remained variable.

These results demonstrate substantial variation in alcohol strength across beverage types, supporting the use of ethanol-standardized measures in subsequent analyses.

### Retail Prices of Alcoholic Beverages

Retail prices were examined to assess nominal differences across beverage categories prior to ethanol standardization.

Retail prices differed markedly across beverage types (Table 3). Licit spirits had the highest mean price (UGX 302,334.51), followed by wines (UGX 129,990.52). Opaque beer (UGX 1,161.29) and illicit spirits (UGX 2,087.63) were substantially cheaper, while clear beer occupied an intermediate position (UGX 7,346.15).

**Table 3:**
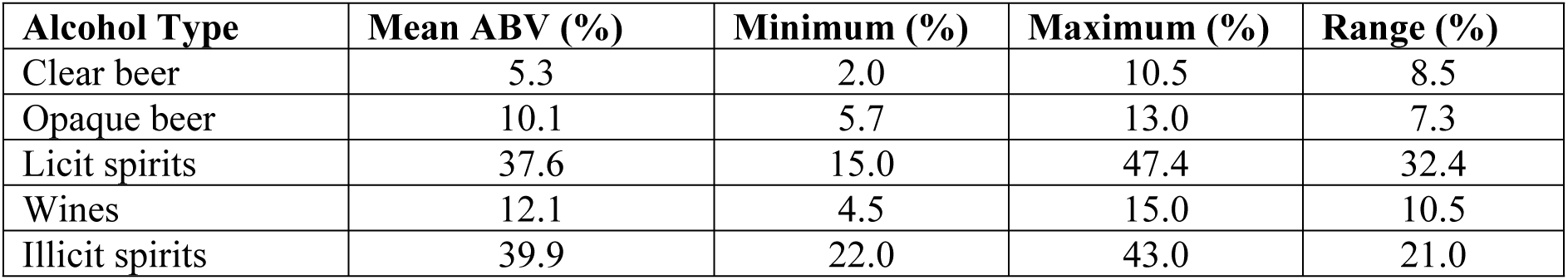
Ethanol Concentration by Beverage Type.

Price dispersion was greatest among licit spirits and wines, as reflected by large standard deviations and wide price ranges. In contrast, opaque beer and illicit spirits exhibited narrower price ranges.

Illicit spirits were characterized by low prices and smaller packaging sizes, resulting in lower per-purchase costs. This price structure suggests the coexistence of higher-cost formal products alongside lower-cost alternatives.

### Cost per Unit of Pure Ethanol

To enable comparability across beverages with different strengths and volumes, prices were standardized into cost per gram of pure ethanol.

Substantial differences in ethanol affordability were observed (Table 4). Opaque beer had the lowest cost per gram of ethanol (UGX 4.25), followed by illicit spirits (UGX 10.49) and clear beer (UGX 19.6). Wines (UGX 170.07) and licit spirits (UGX 375.91) were considerably more expensive.

**Table 4:** Retail Prices of Alcoholic Beverages in Ugx.

| **Category** | **Avg. Vol (ml)** | **Mean Prices** | **Min Prices** | **Max Price** | **Std. Dev.** |
| --- | --- | --- | --- | --- | --- |
| Clear Beer | 389 | 7,346.15 | 3,000.00 | 20,000.00 | 4,624.27 |
| Opaque Beer | 276 | 1,161.29 | 1,000.00 | 4,000.00 | 553.58 |
| Licit Spirits | 737 | 302,334.51 | 5,000.00 | 3,200,000.00 | 472,954.00 |
| Wines | 778 | 129,990.52 | 25,000.00 | 3,300,000.00 | 274,981.40 |
| Illicit Spirits | 200 | 2,087.63 | 1,000.00 | 5,000.00 | 707.15 |

These results indicate that lower-cost beverage categories provide cheaper sources of ethanol relative to formal products. The observed variation highlights the importance of standardized metrics in assessing alcohol affordability.

### Affordability of Intoxication

Marked differences were observed across beverage categories (Table 5). Opaque beer provided the lowest-cost pathway to intoxication (UGX 255), followed by illicit spirits (UGX 629) and clear beer (UGX 1,176). In contrast, wines (UGX 10,204) and licit spirits (UGX 22,555) were substantially more expensive.

**Table 5:** Cost per Gram of Ethanol.

| Product Category | Median Price (UGX) | Mean Ethanol (g) | Cost per gram of ethanol (UGX) |
| --- | --- | --- | --- |
| Clear Beer | 300 | 16.4 | 19.6 |
| Opaque Beer | 101 | 21.7 | 4.25 |
| Licit Spirits | 67,500 | 219.2 | 375.91 |
| Wines | 8505 | 73.8 | 170.07 |
| Illicit Spirits | 800 | 63.0 | 10.49 |

These findings show that the cost of achieving a standardized level of ethanol intake varies widely across beverage types.

### Formal vs Informal Market Comparison

Price variation across market contexts (urban, informal settlements, and rural areas) was examined to assess spatial differences in ethanol pricing.

Clear differences were observed across market contexts (Table 6). Urban markets recorded higher mean ethanol prices across most beverage categories, particularly for licit spirits and wines. Prices declined in urban informal settlements and rural areas.

**Table 6:** Affordability of Intoxication.

| Beverage type | Units needed for intoxication | Cost per gram ethanol (UGX) | Total cost (UGX) | Total cost (USD) |
| --- | --- | --- | --- | --- |
| Clear Beer | 60 | 19.6 | 1,176 | 0.32 |
| Opaque Beer | 60 | 4.25 | 255 | 0.07 |
| Licit Spirits | 60 | 375.91 | 22,555 | 6.08 |
| Wines | 60 | 170.07 | 10,204 | 2.75 |
| Illicit Spirits | 60 | 10.49 | 629 | 0.17 |

1USD = Ugx 3708.57 (Source BoU 08/April/2026)

**Table 7:** Cost per Ml by Alcohol Type and Location.

| Alcohol Type | Urban |  | Urban Slum |  | Rural |  |
| --- | --- | --- | --- | --- | --- | --- |
|  | Mean | Std. Dev | Mean | Std. Dev | Mean | Std. Dev |
| Clear Beer | 32.2 | 11.44 | 13.3 | 5.93 | 10.6 | 2.82 |
| Opaque Beer | - | - | 4.4 | 1.67 | 4.1 | 1.40 |
| Licit Spirits | 551.5 | 651.96 | 83.9 | 58.58 | 53.9 | 19.82 |
| Wines | 190.8 | 393.6 | 52.5 | 20.24 | 46.7 | 13.63 |
| Illicit Spirits | - | - | 11.0 | 3.75 | 10.3 | 3.45 |
| Total | 338.0 | 548.1 | 39.3 | 48.7 | 19.7 | 20.2 |

For example, the cost per millilitre of ethanol for licit spirits decreased from UGX 551.5 in urban areas to UGX 83.9 in informal settlements and UGX 53.9 in rural settings. Similar patterns were observed for clear beer.

Opaque beer and illicit spirits were primarily observed in non-urban markets and were consistently priced at lower levels. Overall average ethanol prices were higher in urban markets (UGX 338.0/ml) compared to informal settlements (UGX 39.3/ml) and rural areas (UGX 19.7/ml).

### Multivariate Analysis of Factors Influencing the Affordability of Ethanol

A multivariate regression model was estimated to identify independent predictors of ethanol affordability while controlling for product, packaging, and market characteristics.

**Table 8:** A Multiple Linear Regression Model Showing Factors Influencing the Affordability of Ethanol.

| Cost Per ml of Pure Alcohol | Coef. | P>z | [95% Conf. Interval] |  |
| --- | --- | --- | --- | --- |
| **Country of Origin** |  |  |  |  |
| Uganda (Ref) |  |  |  |  |
| Imported | 218.6 | **0.00** | 187.21 | 250.03 |
| **Location of Sale (Ref - Urban)** |  |  |  |  |
| Urban Slum | -273.8 | **0.00** | -301.57 | -246.08 |
| Rural | -282.1 | **0.00** | -311.44 | -252.79 |
| **Alcohol Type (Ref – Licit Spirits)** |  |  |  |  |
| Clear Beer | -363.3 | **0.00** | -429.40 | -297.13 |
| Opaque Beer | -508.2 | **0.00** | -661.70 | -354.80 |
| Wines | -79.5 | **0.00** | -121.46 | -37.58 |
| Illicit Spirits | -147.0 | **0.02** | -272.82 | -21.12 |
| **Packaging Type (Ref – Plastics)** |  |  |  |  |
| Can | 183.8 | **0.00** | 73.97 | 293.59 |
| Glass | 95.4 | **0.01** | 19.56 | 171.18 |
| **Retail Outlet Type (Ref – Shop)** |  |  |  |  |
| Bar | 179.2 | **0.00** | 145.92 | 212.52 |
| Kiosk | 59.8 | **0.02** | 11.65 | 107.86 |
| **Packaging Size (Ref - Half Pint (200ml))** |  |  |  |  |
| Half Bottle (375ml) | 127.9 | **0.00** | 64.84 | 190.89 |
| Standard Bottle (750ml) | 19.8 | 0.50 | -37.98 | 77.56 |
| Litres | -7.4 | 0.83 | -76.41 | 61.57 |
| **Availability of the Drink (Ref Monthly)** |  |  |  |  |
| Daily | 7.3 | 0.63 | -22.24 | 36.75 |
| Weekly | -46.3 | **0.00** | -74.40 | -18.10 |
| **Distribution Channel (Ref – Mobile Vendor)** |  |  |  |  |
| Whole saler | 13.8 | 0.52 | -28.05 | 55.62 |
| **Illicitness (Ref – Illicit)** |  |  |  |  |
| Licit | 24.8 | 0.70 | -100.88 | 150.53 |
| **ABV% Content** | 353.5 | **0.00** | 193.25 | 513.66 |

N=823 Observations

The model demonstrated strong explanatory power (Wald χ² significant at p < 0.001; adjusted R² = 0.857), indicating that variation in ethanol cost is largely explained by observed characteristics.

At the product level, imported alcoholic beverages were significantly more expensive than domestically produced ones (β = +196.18 UGX/ml, p < 0.001), implying that intoxication through imported products is less affordable. In contrast, relative to licit spirits, all other beverage categories offered significantly cheaper ethanol, including clear beer (β = −411.64, p < 0.001), opaque beer (β = −622.16, p < 0.001), wines (β = −108.21, p < 0.001), and illicit spirits (β = −200.86, p = 0.001). These differences indicate that the cost of achieving intoxication varies substantially by beverage type, with illicit spirits providing a notably lower-cost source of ethanol.

Packaging characteristics were also associated with the affordability of ethanol. Beverages packaged in glass bottles were significantly more expensive than those in plastic containers (β = +100.55, p = 0.001), indicating that plastic-packaged alcohol represents a cheaper source of ethanol. Packaging size effects were mixed: half bottles (375 ml) were more expensive than half-pints (β = +93.72, p = 0.002), while standard bottles (β = −3.64, p = 0.881) and litre containers (β = −29.82, p = 0.303) did not differ significantly.

The retail environment influenced the cost of intoxication. Alcoholic beverages sold in bars were significantly more expensive than those sold in shops (β = +136.58, p < 0.001), whereas kiosks did not differ significantly from shops (p = 0.228), indicating that the price of ethanol and thus the affordability of intoxication varies across retail settings.

At the market level, substantial geographic differences were observed. Compared to urban centres, beverages sold in urban slums (β = −243.07, p < 0.001) and rural areas (β = −245.22, p < 0.001) were significantly cheaper, indicating that intoxication is more affordable in these settings. Similarly, products sold in low-income (β = −154.91, p < 0.001) and middle-income (β = −175.13, p < 0.001) markets were significantly cheaper than those in high-income areas, demonstrating a clear socioeconomic gradient in the affordability of ethanol.

At the policy level, neither licitness (β = +23.45, p = 0.715) nor alcohol strength (ABV%; β = +135.19, p = 0.204) showed a statistically significant association with ethanol prices after controlling for other factors, suggesting that differences in the affordability of intoxication may not be explained by regulatory status or alcohol strength alone.

Availability and distribution factors were not significantly associated with ethanol prices, including daily (p = 0.347) and weekly availability (p = 0.199), as well as wholesaler distribution (p = 0.680).

## Discussion

This study provides empirical evidence that, in heterogeneous alcohol markets such as those in Uganda, affordability is more accurately conceptualized as access to ethanol rather than beverage price alone. By applying ethanol-standardized pricing and an affordability-of-intoxication framework, the analysis demonstrates that the economic accessibility of alcohol is systematically shaped by the interaction of product type, packaging, and retail context. This extends existing alcohol policy literature by offering a more precise and policy-relevant metric for assessing affordability in low- and middle-income settings.

Consistent with prior research, alcohol affordability is a key determinant of consumption and harm. However, unlike studies in high-income settings where affordability is largely driven by formal pricing structures and standardized products, the present findings show that in fragmented retail environments, variation in alcohol strength and product heterogeneity fundamentally alter how affordability operates. The substantial differences in alcohol by volume (ABV) across beverage types reinforce arguments by (34,36) that ethanol-standardized measures are essential for meaningful comparison. This study advances that position by demonstrating empirically that beverage-level price comparisons alone may underestimate access to pure alcohol in settings where low-cost, high-strength products are widely available.

A central finding is the exceptionally low cost of achieving intoxication through specific beverage categories, particularly opaque beer and illicit spirits. These products provide the most affordable pathways to consuming 60 grams of ethanol, indicating that harmful levels of intake are economically accessible at minimal expenditure. This extends the conceptual work on the “price of intoxication” by (8,23) by showing how such dynamics operate in informal and weakly regulated markets. The persistence of low-cost intoxication reflects underlying structural mechanisms, including low production costs, tax avoidance, and the use of informal supply chains that bypass regulatory controls. This implies that policies targeting formal sector pricing alone may have limited impact where a substantial share of ethanol is supplied through informal channels.

The results further reveal a dual market structure in which high-cost formal products coexist with low-cost informal alternatives. While this pattern has been documented in sub-Saharan Africa (18), this study shows more explicitly how the informal segment actively shapes overall ethanol affordability by sustaining access to cheap alcohol across retail environments. Historically embedded systems of local production, combined with limited enforcement capacity, enable informal products, particularly illicit spirits, to remain widely available and price competitive (19). This implies that formal market expansion does not necessarily displace informal supply, but rather operates alongside it, maintaining structural conditions for low-cost intoxication.

Multivariate analysis provides additional insight into the determinants of ethanol affordability. Product type emerged as the dominant predictor, with all beverage categories offering significantly cheaper ethanol relative to licit spirits. Importantly, illicit spirits remained significantly more affordable even after controlling for packaging and retail factors, indicating that their low price is not solely a function of packaging or point of sale but reflects deeper cost structures within informal production systems plus the absence of revenue taxation. Packaging also played a significant role: alcohol sold in plastic containers was consistently cheaper than that in glass bottles, likely due to lower material (37), transport, and storage costs, as well as the flexibility of informal repackaging practices. These findings suggest that cost-reduction strategies operate across multiple stages of the supply chain.

Geographic and socioeconomic gradients further highlight important equity dimensions. Ethanol was significantly cheaper in rural areas and informal settlements, indicating that lower-income populations face greater exposure to affordable intoxication. This pattern is consistent with evidence linking alcohol affordability to health inequalities and suggests that weaker regulatory oversight, lower operating costs, and the concentration of informal distribution networks contribute to spatial disparities in alcohol access (2,6). This implies that alcohol-related harm may be disproportionately concentrated among economically disadvantaged populations, not because of higher spending, but due to easier access to low-cost ethanol.

Notably, neither alcohol strength (ABV) nor licitness was independently associated with ethanol price after adjustment. This finding suggests that regulatory status alone does not determine affordability and highlights the limitations of policy approaches that focus exclusively on formal sector controls. Instead, market dynamics particularly supply chains, packaging practices, and retail environments appear to play a more decisive role in shaping access to ethanol. This implies that effective policy responses must extend beyond product classification to address the broader structure of alcohol markets.

From a policy perspective, these findings reinforce the importance of ethanol-based regulatory approaches. Pricing interventions, including taxation and minimum unit pricing, are likely to be more effective when explicitly linked to alcohol content (2), thereby directly targeting the affordability of intoxication. However, the persistence of low-cost ethanol in informal markets implies that such measures must be complemented by strengthened regulation of informal production and distribution systems. In addition, the widespread availability of small, low-cost packaging formats suggests a need for policies that address packaging practices that facilitate incremental and affordable consumption.

### Study Strength and Limitations

This study fills an empirical gap by providing a systematic analysis of ethanol affordability and the cost of intoxication in Uganda, combining ethanol-standardized pricing with multivariate regression to identify structural determinants of alcohol access. Including both formal and informal markets, capturing variation in product type, packaging, and retail context, and examining geographic and socio gradients, the study offers novel insights into how low-cost intoxication is distributed across populations and settings.

However, some limitations should be noted. The cross-sectional design precludes causal inference, and retail price observations do not necessarily reflect actual purchasing or drinking behaviour. Measurement error in alcohol content may persist, particularly for informal and illicit products, due to variability in production processes. The study only included alcohol brands with reported ABV% and observable features like Q-Marks and revenue stamps; however, the majority of alcohol in Uganda is home-brewed. Furthermore, the study did not undertake laboratory testing to determine what was contained in the various alcohol products. Thus, a feature laboratory investigation on what is contained in alcohol is necessary in order to determine the illicitness of alcohol brands and identify illicit brands that disguise as licit. Additionally, the absence of individual-level income or expenditure data limits the ability to assess affordability relative to personal purchasing power. Despite these constraints, the study provides robust, policy-relevant evidence on economic access to ethanol in a heterogeneous alcohol market.

## Conclusion

The study revealed that in Uganda’s fragmented alcohol market, alcohol affordability is largely determined by access to ethanol rather than the price of specific alcoholic beverages. The findings showed that harmful intoxication can be achieved at a very low cost through informal and inexpensive alcohol products, making excessive consumption economically accessible to many individuals. This suggests that beverage-based alcohol control policies alone may be insufficient to reduce alcohol-related harm. Therefore, governments and stakeholders should adopt ethanol-based regulatory measures such as minimum unit pricing and alcohol content adjusted taxation, strengthen the regulation and monitoring of informal alcohol markets, and target interventions in rural and low-income communities where cheap alcohol is most accessible. In addition, national monitoring systems should incorporate ethanol-standardized affordability indicators, while future research should examine the relationship between alcohol affordability, consumption patterns, and health outcomes to support more effective alcohol control strategies.

## Acknowledgements

We acknowledge the support of local field teams and retail vendors who provided access to pricing data, and the administrative staff of Arrow Empirical Research and Skills Enhancement Co. Ltd. (AERSE) for their assistance in data management.

## Author Contributions

All nine authors contributed to the study as follows: conceptualization and design (MT, SPR, KN); data collection and curation (SPR, KN); analysis and interpretation (SPR, MT); manuscript drafting (SPR, MT); BK: Formal analysis, Reviewing & editing, (CZ, TRM): Editing and reviewing. AR: Funding acquisition and Resources, Conceptualization and review & editing. TA: Conceptual development, funding acquisition, investigation, methodological design, project administration, resource mobilization, and manuscript preparation, including review and editing: Critical revision and approval of final manuscript (all authors All authors take responsibility for the content of the manuscript.

## Data Availability Statement

Data supporting the findings of this study is available from the corresponding author upon reasonable request, subject to privacy and ethical considerations.

## Abbreviations

ABV: Alcohol by Volume
UGX: Ugandan Shilling
WHO: World Health Organization
LMIC: Low- and Middle-Income Countries

